# Meta-analysis of coagulation disbalances in COVID-19: 41 studies and 17601 patients

**DOI:** 10.1101/2021.10.17.21265108

**Authors:** Polina Len, Gaukhar Iskakova, Zarina Sautbayeva, Aigul Kussanova, Ainur T. Tauekelova, Madina M. Sugralimova, Anar S. Dautbaeva, Meruert M. Abdieva, Eugene D. Ponomarev, Alexander Tikhonov, Makhabbat S. Bekbossynova, Natasha S. Barteneva

## Abstract

**Introduction:** Coagulation parameters are important determinants for COVID-19 infection. We conducted meta-analysis to assess the early hemostatic parameters in retrospective studies in association with severity of infection.

**Methods:** Ovid, PubMed, Web of Sciences, and Google Scholar were searched for research articles that addressed clinical characteristics of COVID-19 patients and disease severity. Results were filtered using exclusion and inclusion criteria and then pooled into a meta-analysis to estimate the standardized mean difference with 95% CI for each of five coagulation parameters (D-dimers, fibrinogen, prothrombin time, platelets count, activated partial thromboplastin time). Two authors independently extracted data and assessed study quality. To explore the heterogeneity and robustness of our fundings, sensitivity and subgroup analyses were conducted. Publication bias was assessed with contour-enhanced funnel plots and Egger test by linear regression.

**Results:** Overall, 41 original studies (17601 patients) on SARS-CoV2 were included. For the two groups of patients, stratified by severity, we identified that D-dimers, fibrinogen, activated partial thromboplastin time, and prothrombin time were significantly higher in the severe group (SMD 0.6985 with 95%CI [0.5155; 0.8815]); SMD 0.661with 95%CI [0.3387; 0.9833]; SMD 0.2683 with 95%CI [0.1357; 0.4009]; SMD 0.284 with 95%CI [0.1472; 0.4208]). In contrast, PLT was significantly lower in patients with more severe cases of COVID-19 (SMD −0.1684 with 95%CI [-0.2826; −0.0542]). Neither the analysis by the leave-one-out method nor the influence diagnostic have identified studies that solely cause significant change in the effect size estimates. Subgroup analysis showed no significant difference between articles originated from different countries but revealed that severity assessment criteria might have influence over estimated effect sizes for platelets and D-dimers. Contour-enhanced funnel plots and the Egger test for D-dimers and fibrinogen revealed significant asymmetry that might be a sign of publication bias.

**Conclusions:** The standard coagulation laboratory parameters with exception of platelets counts are significantly elevated in patients with severe COVID-19. However, fibrinolysis shutdown requires evaluation outside conventional coagulation tests and analysis of additional specific markers related to clotting formation and PLT characteristics. We hypothesize that a proportion and parameters of immature reticulated platelets may serve as additional biomarkers for prediction of adverse events.

## 1 Introduction

In less than two years of the outbreak, the COVID-19 pandemic took almost five million lives (John Hopkins, 2021). SARS-CoV-2 is the seventh member of the large coronavirus family capable of inducing human disease (Carsana et al., 2020). Viral respiratory infections, including severe respiratory acute syndrome coronavirus (SARS-CoV), Middle East respiratory syndrome coronavirus (MERS-CoV), SARS-CoV-2 may induce coagulopathy and lead to intravascular thrombi and deposition of fibrinogen (Hwang et al., 2005; Yang et al., 2005; Goeijenbier et al., 2012; WHO MERS guidelines, 2013; Giannis et al., 2020). The coagulation system is activated and dysregulated during COVID-19 infection; however, characteristics of COVID-19 associated coagulopathy are different from coagulation disorders (Iba et al., 2020). Evidence of abnormal COVID-19-associated coagulation parameters appeared already in early publications from Wuhan (Chen et al., 2020; Wang et al., 2020; Zhou et al., 2020) and was supported by further publications worldwide (rev. Levi et al., 2020; Iba et al., 2020). The reported hemostatic abnormalities with COVID-19 infection include increased D-dimers levels (Lippi et al., 2020; Demelo-Rodriguez et al., 2020), fibrinogen levels (Ranucci et al., 2020; Panigada et al., 2020), changes in the quantities and levels of activation of platelets (Yang et al., 2020; Lippi et al., 2020; Hottz et al., 2020; Althaus et al., 2021), elevated von Willebrand factor (Goshua et al., 2020; Doevelaar et al., 2021, rev. by Becker et al., 2021) and other coagulation parameters. Persistently elevated thrombocytic events, even with initiation of prophylactic anti-coagulation, suggest the presence of hypofibrinolysis in addition to detected hypercoagulability in COVID-19 (Creel-Bulos et al., 2021; Bachler et al., 2021). It is also consistent with results of thromboelastography (TEG) reported by Wright et al. (2020).

There is a need in early detection of elevated coagulation biomarkers to optimize risk stratification of patients with COVID-19. However, due to limitations of early pandemic research with small study sizes and highly heterogeneous datasets it is still debated. The aim of the current study was to evaluate the validity of stratification based on early coagulation parameters provided by different clinical studies worldwide. We also aimed to use subgroup analysis to explore whether heterogeneity in results was in part explained by differences in patient subpopulations. Finally, the coagulation parameters of our own cohort were included in a systematic meta-analysis of published studies.

## 2 Materials and Methods

In this study, we aimed to identify the relationship between coagulation biomarkers taken at admission and severity of COVID-19 in adult (>18) patients by estimating the effect size of five laboratory coagulation tests: D-dimers, platelets, fibrinogen, activated partial thromboplastin time, and prothrombin time.

### 2.1 Search Strategy, Exclusion and Inclusion criteria

To broaden our search, we included Ovid, PubMed, Web of Sciences, and Google Scholar in the study. They were thoroughly scanned using the following keywords: SARS-CoV-2, COVID-19, coagulation, severity, characteristics, features, D-dimers, platelets, fibrinogen, cohort, observational, retrospective. First, satisfying search results were exported to an Excel table and duplicates were eliminated. Second, we conducted a preliminary review of retrieved articles’ abstracts. Papers that focused on pregnant women, children, or specific age groups were avoided. Similarly, works that focused on the effect of medical treatment or patients with particular commodities were not included in the meta-analysis. We also excluded studies that compared mortality among groups of COVID-19 patients. Then, each publication was checked according to the inclusion criteria: sample size, severity assessment criteria, presence of at least one coagulation marker of interest expressed as a continuous variable. To be included, a study should have had more than 15 patients and divided them according to the disease severity - WHO guidelines, ICU admission, disease aggravation, or need for oxygen therapy. Next, a list of biomarkers should have included at least some of the coagulation parameters: platelets count (PLT), D-dimers (DD), fibrinogen (FIB), activated partial thromboplastin time (APTT), prothrombin time (PT), thrombin time (TT), Activated thrombin (AT), Factor VIII (FVIII), and Von Willebrand factor (vWF). Finally, the quality of selected articles was assessed with the NIH Quality Assessment Tool for Observational Cohort and Cross-Sectional studies. Questions 5, 6, 12, 13 were not applicable to selected studies due to their observational/retrospective nature; Question 8 was not applicable to studies with dichotomous exposure values, like ICU admission and need for oxygen therapy. Hence, studies with a quality score of less than five were excluded from the analysis. Search, and quality check were conducted independently by two authors. Disagreements were resolved during a joint discussion with a third author.

### 2.2 Data extraction

The primary goal was to retrieve from each article data about two groups of patients differing by disease severity: group size, stratification criteria, results of the laboratory tests. Apart from these, we kept a record of the country, hospital, and admission time in order to identify and exclude duplicate patients.

### 2.3 Cohort from the National Research Center for Cardiac Surgery

#### 2.3.1 Participants and data collection

From a large cohort of 560 patients consecutively admitted from June 2020 to August 2021 due to coronavirus infection to the cardiology department of the NCJSC “National Research Center for Cardiac Surgery” (NRCCS), Nur-Sultan, Kazakhstan, we selected 451 patients with a confirmed diagnosis of COVID-19, aged ≥18 years, and those with the onset of the disease ≤ 21 days, all of whom were hospitalized for at least 24 hours with COVID-19. All demographic, clinical, laboratory data were extracted from the electronic records of the NRCSC. Laboratory tests included PLT, PT, international normalized ratio, APTT, FIB, DD. Blood samples were collected at admission by the clinical team. Clotting tests were performed according to standard methods. For coagulation tests (PT, international normalized ratio, APTT, FIB), we used AVATUBE® with sodium citrate, and it was measured on Sysmex CS-2500 (Sysmex, Japan). Blood for DD was collected in AVATUBE® with sodium citrate, and D-dimers were quantified on Cobas® 6000. PLT counts were measured on Sysmex XS-500 (Sysmex, Japan) with blood collection performed in AVATUBE® with K2 EDTA. The confirmation of SARS-CoV-2 infection was done by real-time quantitative reverse-transcription polymerase chain reaction (RT-PCR) assay on nose/throat swab or sputum samples using CFX96® Real-Time System (Bio-Rad Laboratories, Inc., USA).

#### 2.3.2 Clinical assessment

The COVID-19 population was initially divided into four groups according to disease severity following local guidelines “Diagnosis and treatment of Covid-19 in Adults” (http://www.rcrz.kz/index.php/ru/2017-03-12-10-51-13/klinicheskie-protokoly) (**Suppl. File 1**). The study protocol was approved by the medical ethics committee of the National Research Center for Cardiac Surgery, Nur-Sultan, Kazakhstan waiving the need for written informed consent due to the retrospective design.

### 2.4 Data analysis

#### 2.4.1 Data transformation

All data analysis was performed in the RStudio 1.4.1717 with R version 4.1.0 (Integrated Development Environment for R. RStudio, PBC, USA). In the most of cases, results were reported as the first, second (median), and third quartiles, reflecting that original data was not normally distributed. The “estmeansd” package was used to convert median, interquartile range, minimum and maximum values to mean and standard deviation. The package implements Box Cox transformation to normalize raw data prior to conversion. Then, Lou (2018) formulas for mean and Wan (2014) formulas for standard deviation are used. Since the criteria and number of groups differed from study to study, patients had to be regrouped into the more and less severe groups accordingly: severe/critical and mild/moderate, ICU admitted patients and outpatients/general ward patients, observed aggravation and recovery/stable, and need for oxygen therapy.

#### 2.4.2 Meta-analysis

The “meta” and “metafor” packages were used for the meta-analysis. Heterogeneity was assessed using the I^2^, tau-squared, and prediction intervals. Following the recommendations of Veroniki (2016), Paule-Mandel (PM) tau-squared estimator and Q-Profile (QP) tau-squared confidence interval estimator were used in this meta-analysis. To account for different measurement scales and possible inaccuracy of the Graph Digitizer, the standardized mean difference (SMD), or Hedges g, based on the inverse-variance approach, was estimated. For datasets with high heterogeneity, the Knapp-Hartung adjusted random-effects model is implemented. In case of low heterogeneity, the fixed-effects model is applied.

#### 2.4.3 Sensitivity and Subgroup analysis

We also performed several sensitivity analyses using the “dmetar” package. First, we recalculated the effect size using the leave-one-out method. Second, basic outliers, studies whose confidence intervals do not overlap with the pooled effect confidence interval, were removed. Third, we ran the diagnostic for influential cases with Graphic Display of Study Heterogeneity (GOSH) plots and excluded them from the meta-analysis. To further explore the heterogeneity of the data, we conducted subgroup analyses: based on grouping criteria and location.

### 2.5 Publication Bias

We used the counter-enhanced Funnel plots and Egger’s test by linear regression to assess the possibility for publication bias in selected studies.

## 3 Results

### 3.1 Results of the Web Search

The stepwise process of selection is depicted in Figure 1. Initially, there were 4056 papers that matched search parameters. After the removal of duplicates and studies that did not meet the exclusion criteria, only 195 unique studies remained. A more thorough review based on the inclusion criteria has proven 125 studies to be unfitting. In addition to 70 that were left, 15 studies were extracted from the references lists of previously published meta-analyses and systematic reviews. Finally, we performed a quality check and removed duplicate patients, which resulted in 40 being included into the meta-analysis. Besides original articles, we have included data from the National Research Center for Cardiac Surgery located in Nur-Sultan, Kazakhstan. This data set included data from 451 patients with acute COVID-19.

**Figure 1.**
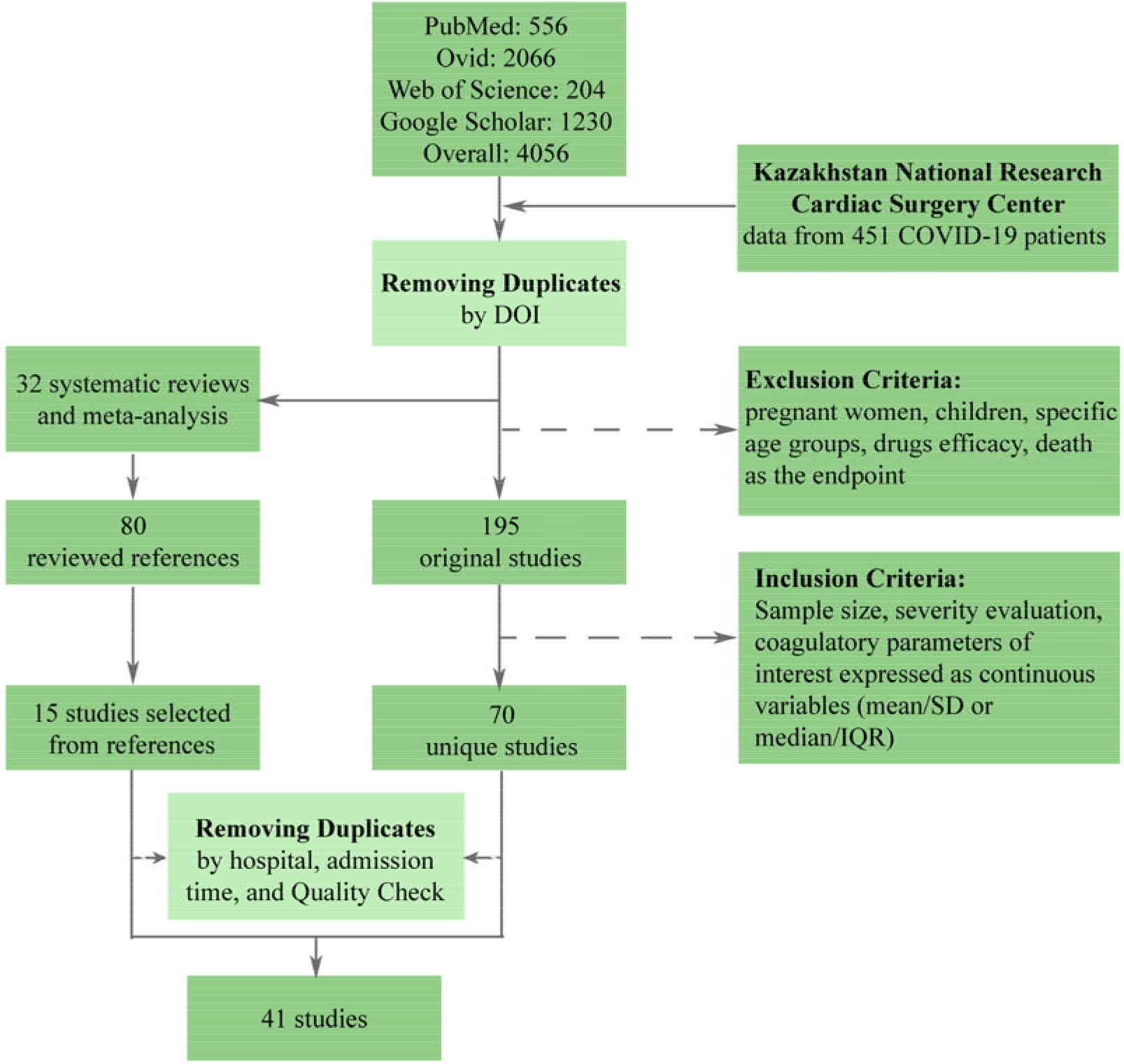
Flow diagram illustrating the process of data collection.

### 3.2 Description of Selected Studies

Overall, this meta-analysis comprises 17601 patients from 40 research papers and the NRCSC. Of the total, 24 studies originated from China, while the rest included data from France, Germany, Italy, Mexico, Singapore, South Korea, Turkey, UK, and the USA. Most of the articles (23) reported division of patients according to the WHO guidelines. Out of the rest, 12 articles made groups based on the ICU admission, 3 focused on disease aggravation, and 3 on the need for oxygen therapy. This information is depicted in **Figure 2** in more detail.

**Figure 2.**
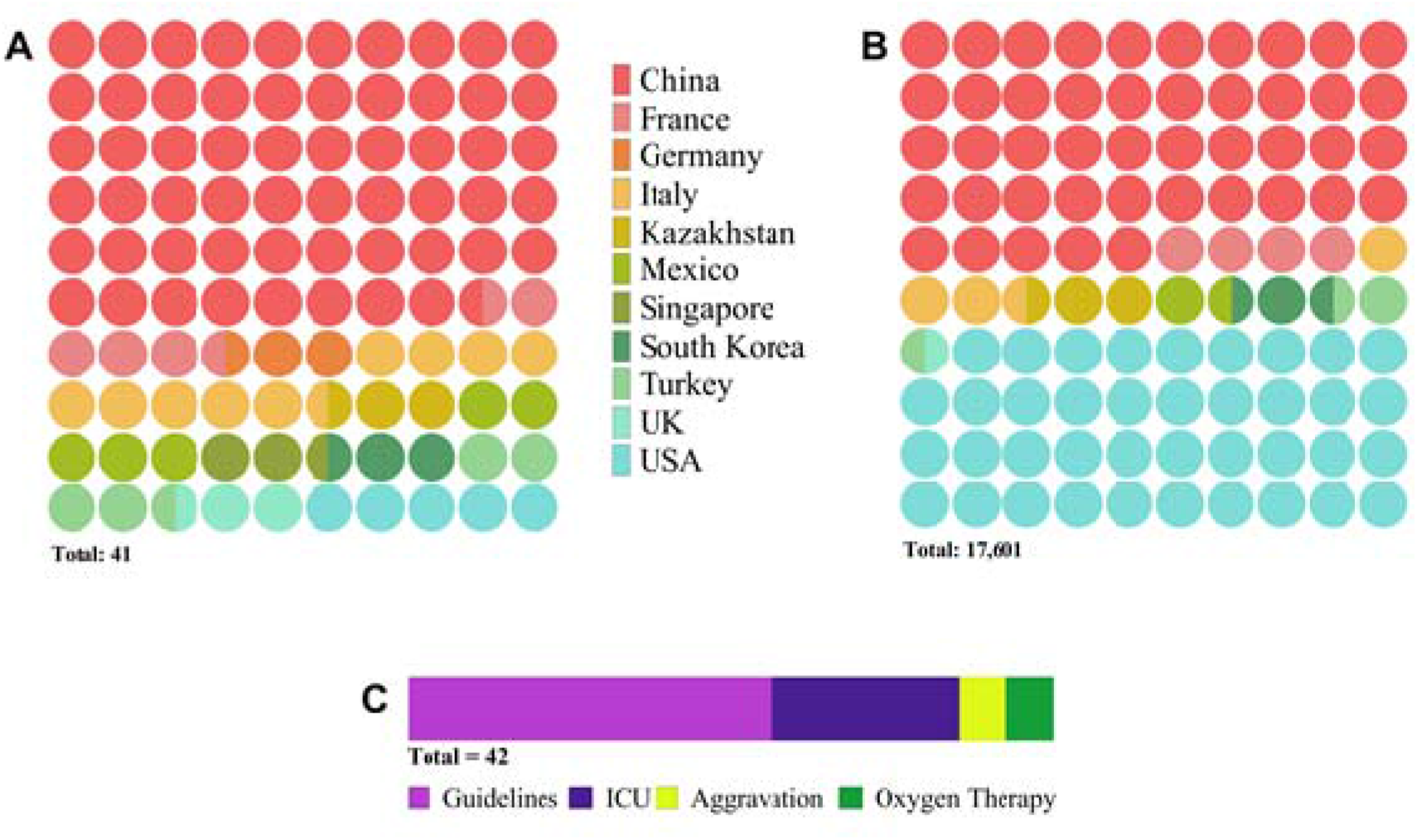
Diagram depicting characteristics of selected studies: (**A**) number of publications per country, (**B**) number of patients per country, (**C**) number of publications per severity assessment criterion. The latter (**C**) indicates the total number of research articles to be 42 instead of 41 because one study reported two populations of patients admitted to the hospital at different periods.

### 3.3 Results of Meta-analysis

#### 3.3.1 Pooled Effect Sizes

We collected data for five coagulation parameters: PLT, DD, FIB, APTT, and PT. In all cases, the heterogeneity was extremely high (I^2^ > 80%), so the Knapp-Hartung adjusted random-effects model was applied in all parts of the meta-analysis. DD, FIB, APTT, and PT were significantly higher in more severe cases (SMD 0.6985 with 95%CI [0.5155; 0.8815]; SMD 0.661 with 95%CI [0.3387; 0.9833]; SMD 0.2683 with 95%CI [0.1357; 0.4009]; SMD 0.284 with 95%CI [0.1472; 0.4208]). In contrast, PLT was significantly lower in patients with severe cases of COVID-19 (SMD −0.1684 with 95%CI [−0.2826; −0.0542]). Forest plots for the meta-analyses are shown in **Figures 3-4**, and results are highlighted in **Table 1**.

**Table 1.** Summary of the effect sizes for all coagulation parameters

| Parameter | Effect Size |  |  |  | Heterogeneity |  |  |
| --- | --- | --- | --- | --- | --- | --- | --- |
|  | Pooled SMD | 95% CI | t | p value | I<sup>2</sup> (%) | tau-squared | p value |
| PLT | -0.1684 | [-0.2826; -0.0542] | -3 | 0.0051 | 82.2 | 0.0743 | <0.0001 |
| DD | 0.6985 | [0.5155; 0.8815] | 7.74 | <0.0001 | 94.2 | 0.2636 | <0.0001 |
| FIB | 0.661 | [0.3387; 0.9833] | 4.27 | 0.0003 | 90.4 | 0.4812 | <0.0001 |
| APTT | 0.2683 | [0.1357; 0.4009] | 4.22 | 0.0004 | 82.8 | 0.0558 | <0.0001 |
| PT | 0.284 | [0.1472; 0.4208] | 4.3 | 0.0003 | 80 | 0.0785 | <0.0001 |

**Figure 3.**
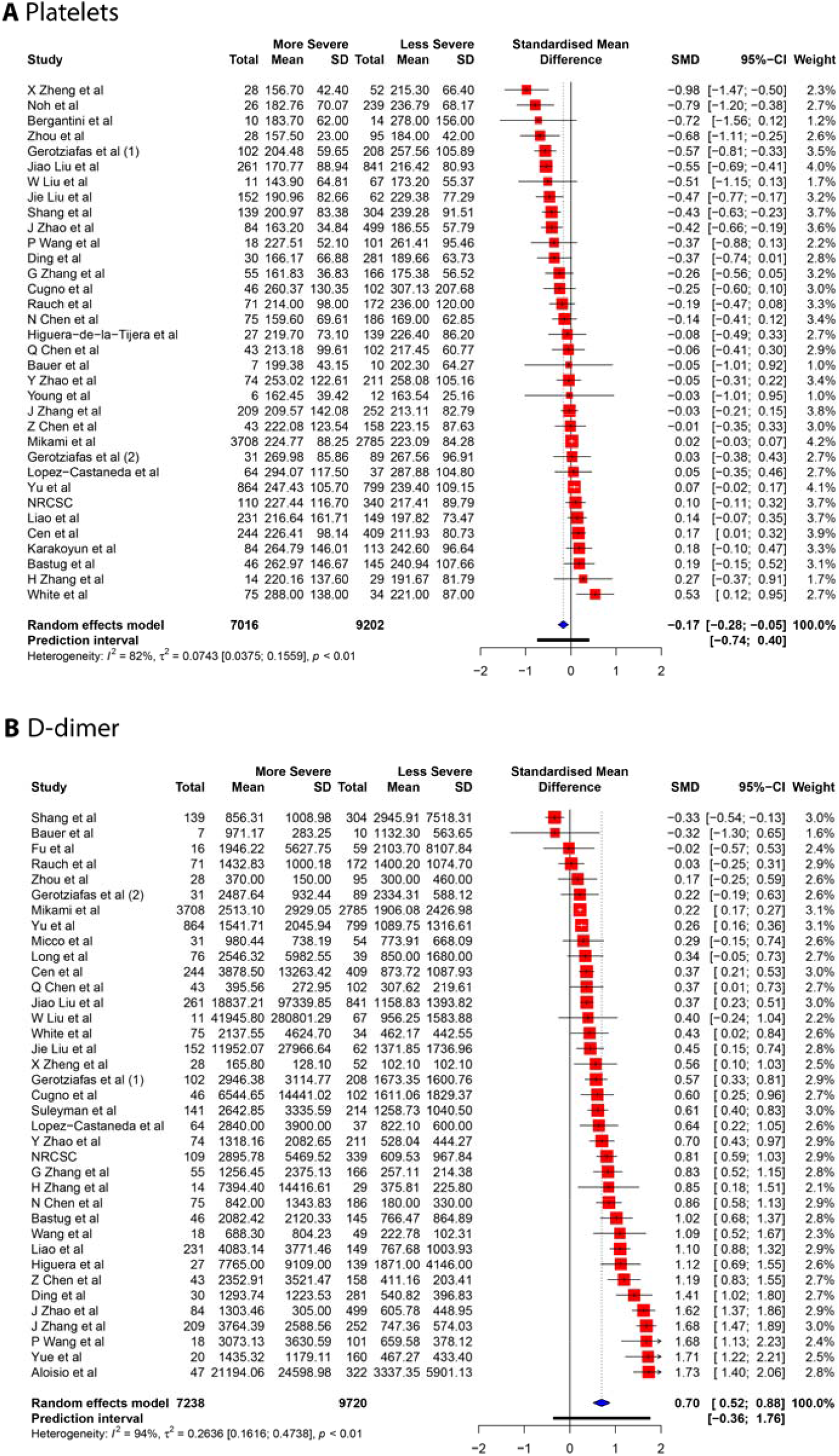
Forest plot of association between COVID-19 severity and platelets (**A**), D-dimers (**B**).

**Figure 4.**
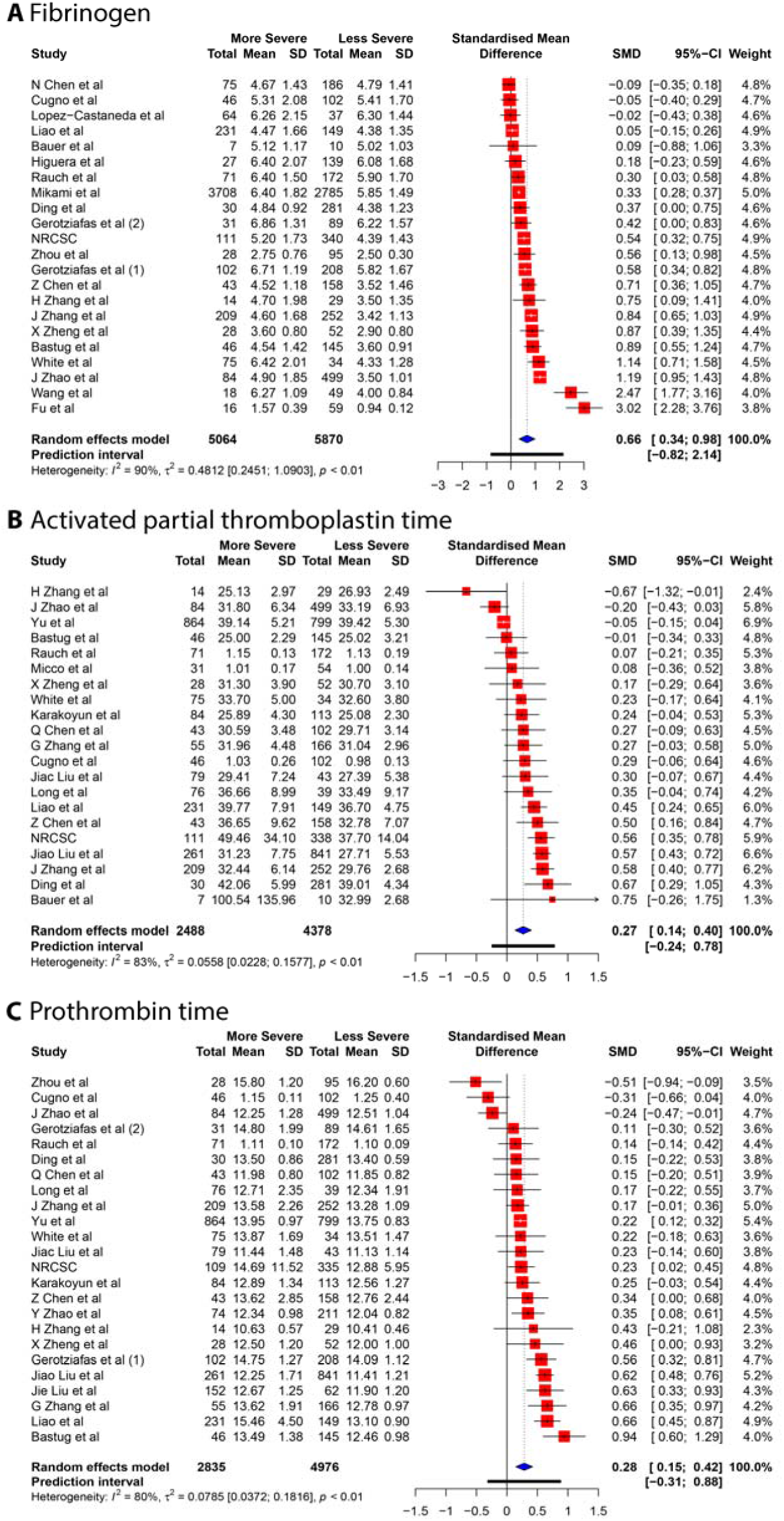
Forest plots of the association between COVID-19 severity and: (**A)** fibrinogen, (**B)** activated partial thromboplastin time, (**C)** prothrombin time.

#### 3.3.2 Sensitivity and Subgroup Analysis

Neither the analysis by the leave-one-out method nor the influence diagnostic has identified studies that solely cause a significant change in the effect size estimates. Removal of several outliers based on the pooled and individual confidence intervals allowed significant decrease in heterogeneity, yet it did not affect the interpretation of the results. Adjusted effect sizes and heterogeneity measurements are summarized in **Table 2**. All influence diagnostic plots, such as Baujat and leave-one-out plots, Cook’s distance, Covariance Ratio, etc., are provided in the **Suppl. Figures 1-16** along with resulting Forest plots.

**Table 2.** Summary results of the sensitivity analysis

| Coagulation Parameter |  | Effect Size |  |  |  | Heterogeneity |  |  |
| --- | --- | --- | --- | --- | --- | --- | --- | --- |
|  |  | Pooled SMD | 95% CI | t | p value | I<sup>2</sup> (%) | tau-squared | Q test p-value |
| PLT | Original | -0.1684 | [-0.2826; -0.0542] | -3 | 0.0051 | 82.2 | 0.0743 | <0.0001 |
|  | Outliers removed | -0.1416 | [-0.2419; -0.0412] | -2.9 | 0.0076 | 56.3 | 0.0287 | 0.0002 |
|  | Inf. Stud. removed | - | - | - | - | - | - | - |
| DD | Original | 0.6985 | [0.5155; 0.8815] | 7.74 | <0.0001 | 94.2 | 0.2636 | <0.0001 |
|  | Outliers removed | 0.621 | [0.4860; 0.7560] | 9.47 | < 0.0001 | 70.2 | 0.073 | < 0.0001 |
|  | Inf. Stud. removed | - | - | - | - | - | - | - |
| FIB | Original | 0.661 | [0.3387; 0.9833] | 4.27 | 0.0003 | 90.4 | 0.4812 | <0.0001 |
|  | Outliers removed | 0.5889 | [0.4141; 0.7638] | 7.14 | < 0.0001 | 85 | 0.0801 | < 0.0001 |
|  | Inf. Stud. removed | 0.4855 | [0.3030; 0.6679] | 5.57 | < 0.0001 | 86.1 | 0.1175 | < 0.0001 |
| APTT | Original | 0.2683 | [0.1357; 0.4009] | 4.22 | 0.0004 | 82.8 | 0.0558 | <0.0001 |
|  | Outliers removed | 0.348 | [0.2450; 0.4518] | 7.14 | < 0.0001 | 38.6 | 0.0125 | 0.0532 |
|  | Inf. Stud. removed | 0.3214 | [0.2036; 0.4392] | 5.73 | < 0.0001 | 68.8 | 0.0326 | <0.0001 |
| PT | Original | 0.284 | [0.1472; 0.4208] | 4.3 | 0.0003 | 80 | 0.0785 | <0.0001 |
|  | Outliers removed | 0.293 | [0.2099; 0.3761] | 7.44 | < 0.0001 | 26.9 | 0.0064 | 0.1407 |

GOSH plots were built for all five models. K-means, DBSCAN, and Gaussian Mixture Model were implemented as clustering methods. This diagnostic was not applied on PLT and DD since they only have one cluster each (**Figure 5**); for FIB, APTT, and PT, on the other hand, we constructed the plots and identified several possible influential cases. Original GOSH plots for these parameters are given at **Suppl. Figures**. For fibrinogen, models that include studies by Fu et al. (2020) and Wang et al. (2020) tend to show higher heterogeneity and greater effect size (**Figure 6A-B**). For activated partial thromboplastin time, GOSH plot demonstrated that studies by J. Zhao et al. (2021), H. Zhang et al (2020), and Yu et al. (2020) increase model’s heterogeneity and slightly pull the effect size to the left (**Figure 6C-E**). GOSH plots of the prothrombin time effect size identified four studies, three of which (J. Zhao et al (2021), Cugno et al. (2021), and Zhou et al. (2021) pull effect size to the left (**Figure 6F-H**), and one (Bastug et al.) pulls effect size to the right (**Figure 6I**). We’ve excluded these studies and recalculated the pooled effect size for each parameter. Results are summarized in **Table 2**.

**Figure 5.**
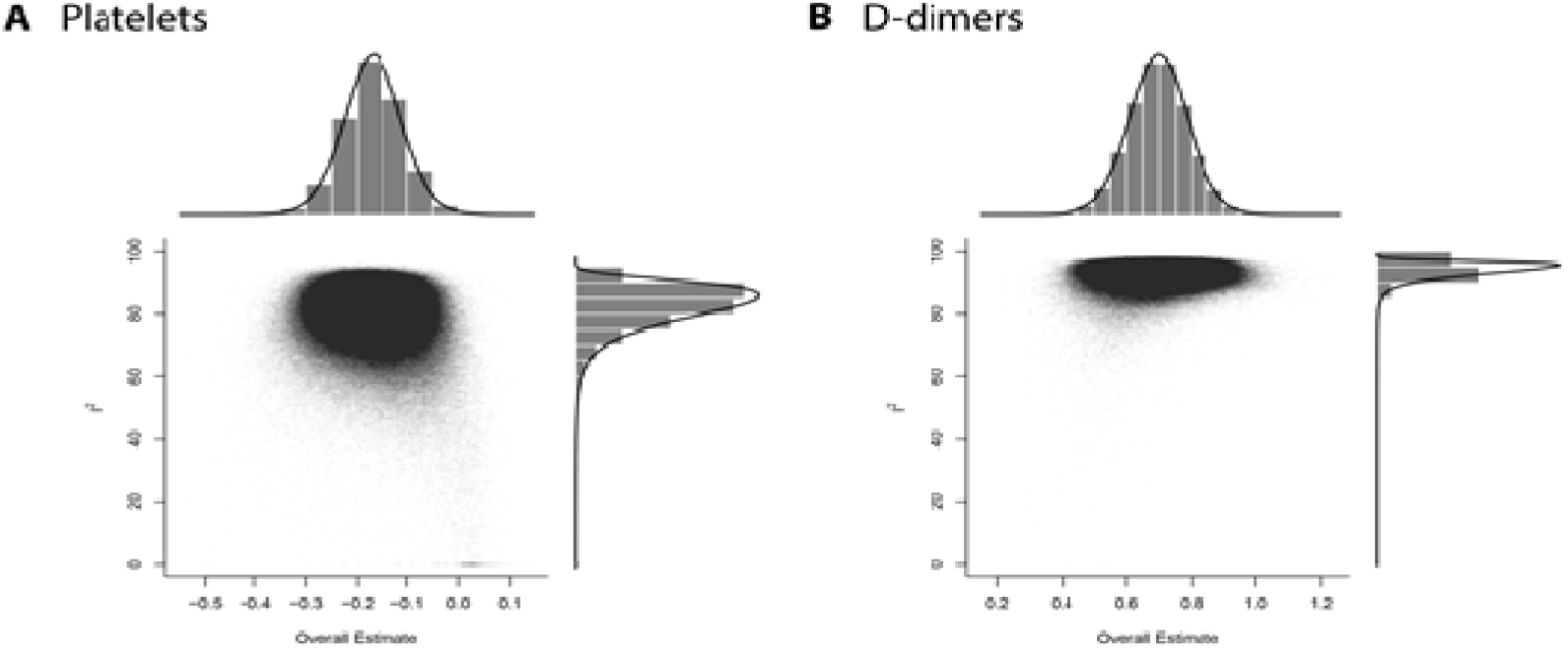
GOSH diagnostic for influential cases in meta-analysis models (**A) -** PLT, (**B) -** DD.

**Figure 6.**
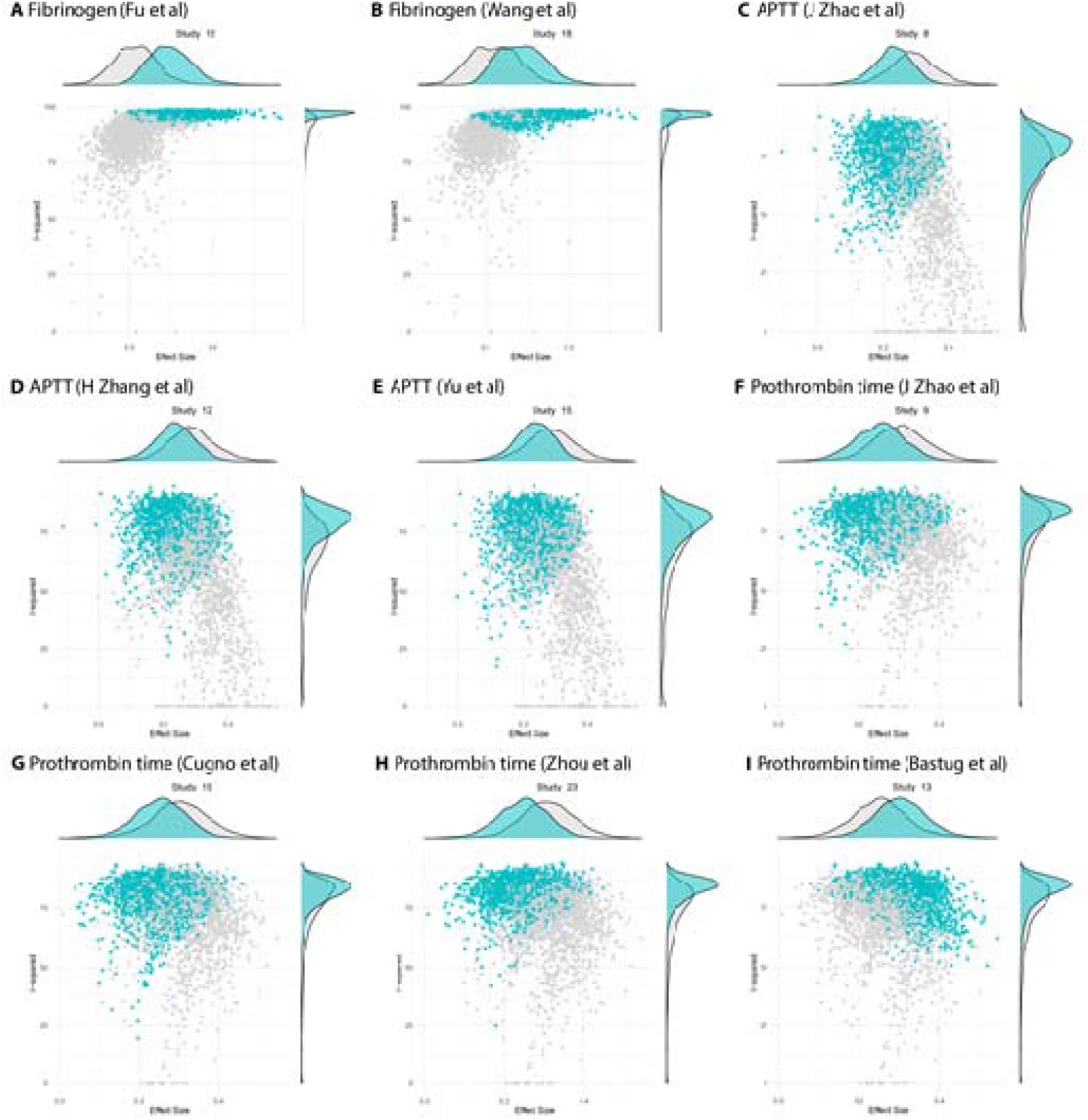
GOSH plots fixed for influential studies in meta-analysis models. (**A-B)** - fibrinogen; (**C-E)** - APTT; (**F-I)** - prothrombin time.

To explore the possible sources of heterogeneity and test the robustness of our data, we have conducted subgroup analyses based on articles’ country of origin and criteria for severity stratification. Results are summarized in **Table 3**. Subgroup analysis showed no significant difference between articles originated from different countries but revealed that severity assessment criteria might have an influence on overestimated effect sizes for PLT and DD. High heterogeneity levels seem to be persistent oven different subgroups. Being the only exception, values of APTT obtained from non-Chinese articles were less heterogenous - 47.1% compared to 84.7% in studies originated in China.

**Table 3.**
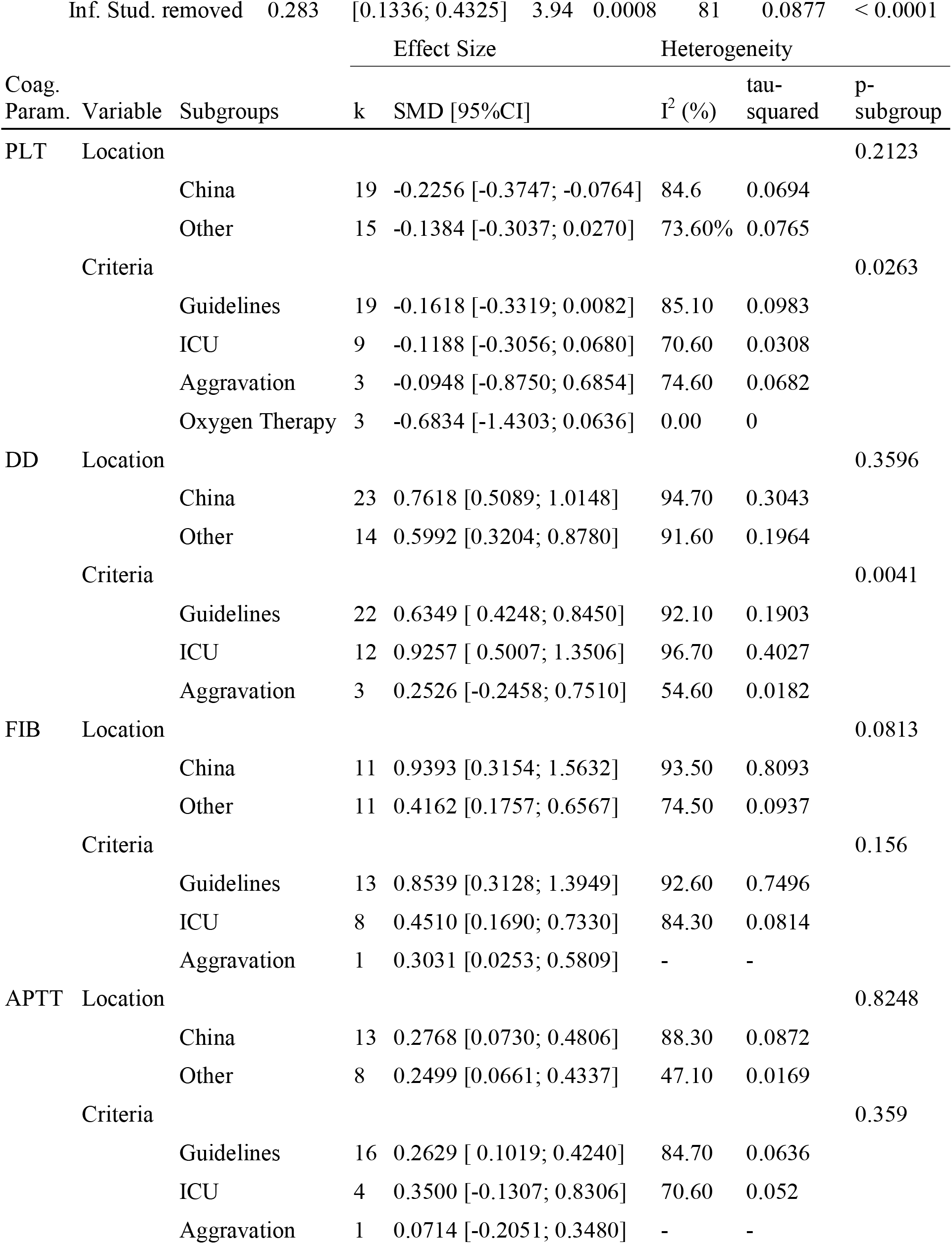
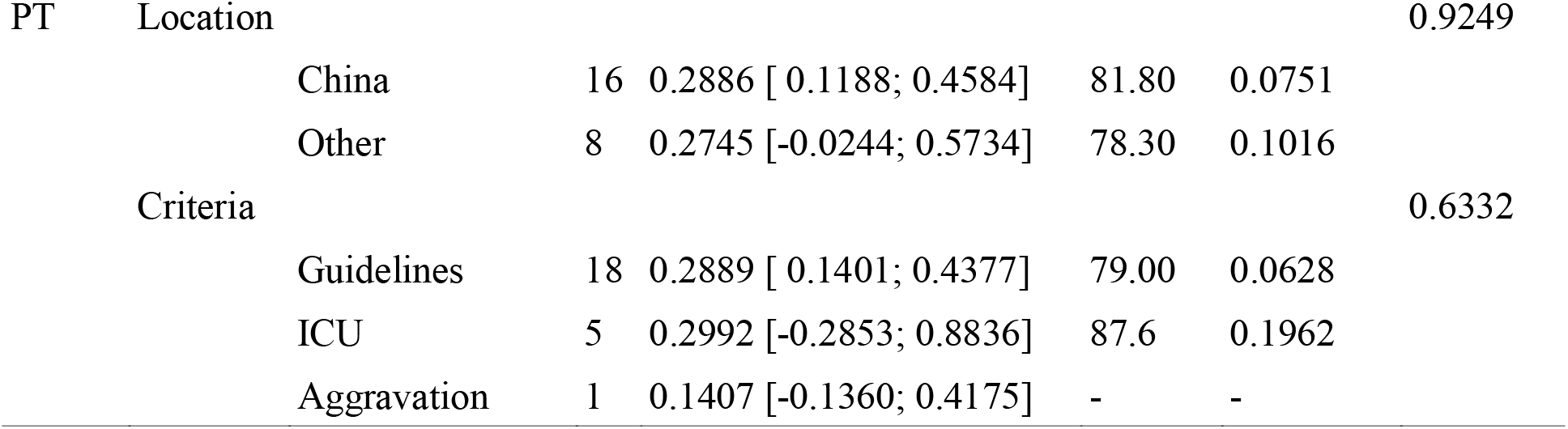
Summary results of the subgroup analysis

#### 3.3.3 Publication Bias

Contour-enhanced funnel plots for five coagulation parameters are depicted in **Figure 7**, and the results of Egger’s test are summarized in **Table 4**. Based on the linear regression results, significant asymmetry is observed in published studies that included results for PLT, DD (Intercept [p-value]: −1.297 [0.0336], 3.182 [0.0013]), and somewhat significant asymmetry was indicated for FIB (1.872 [0.0493]). While it is impossible to predict whether asymmetry is coming solely from publication bias, contour-enhanced funnel plots for DD and FIB (**Figure 6B-C**) show that the number of papers with insignificant findings is very small (p > 0.1). This may advocate for the presence of some publication bias. In contrast, **Figure 6A** illustrates that even though the funnel plot for PLT has some asymmetry, there are plenty of publications with insignificant SMDs, thus decreasing the possibility of publication bias.

**Table 4.** Egger’s test for publication bias

| Parameter | Intercept | Confidence Interval | t | p-value |
| --- | --- | --- | --- | --- |
| PLT | -1.297 | [-2.44, -0.15] | -2.221 | 0.0336 |
| DD | 3.182 | [ 1.39, 4.97] | 3.489 | 0.0013 |
| FIB | 1.872 | [0.12, 3.63] | 2.093 | 0.0493 |
| APTT | 0.775 | [-1.37, 2.92] | 0.708 | 0.4874 |
| PT | -0.436 | [-2.49, 1.62] | -0.417 | 0.6811 |

**Figure 7.**
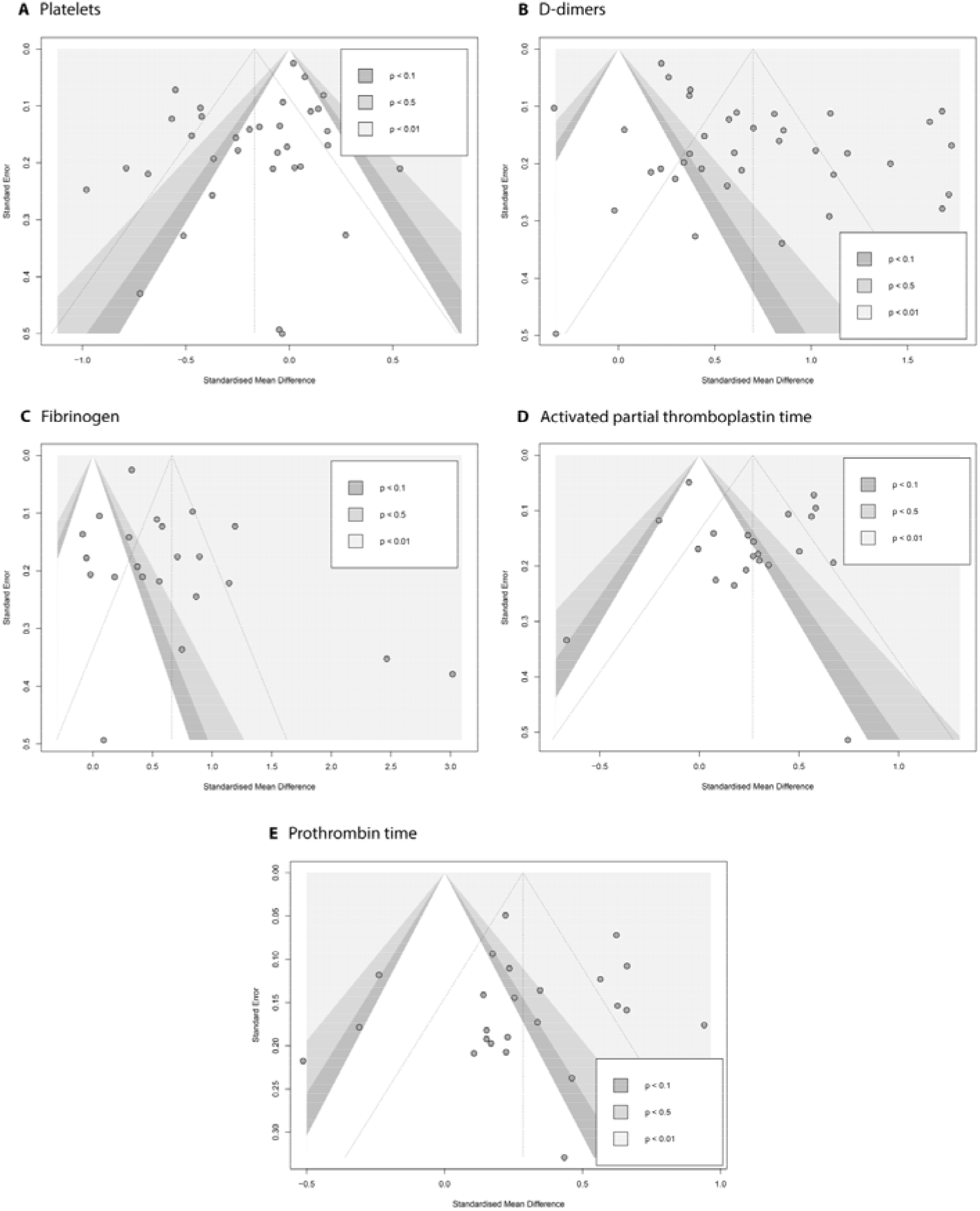
Contour-enhanced funnel plots evaluating the presence of publication bias in the pool of articles that report association between COVID-19 severity and (**A)** platelets; (**B)** D-dimer; (**C)** fibrinogen; (**D)** activated partial thromboplastin time; (**E)** prothrombin time.

To further explore the robustness of our findings, we have conducted several additional tests. As reported by Zwetsloot (2017) and his colleagues, plotting SMD against its standard error to assess the publication bias can result in distorted funnel plots, leading to false-positive conclusions. Since SMD is necessary for our analysis and cannot be substituted, we used a sample size-based precision estimate instead of the standard error – 1/√n. The resulting funnel plots (**Supplemental Figure 16**) showed no significant distortion.

Similar corrections were made to Egger’s test: since standardized mean difference and standard error are not independent, we can expect the test to yield some false-positive results. In our calculations, we used the corrected SE suggested by Pustejovsky and Rodgers (2019). Results are given in the **Supplemental Table 2**. The only model that was affected by this correction was FIB (intercept 2.142 with a p-value of 0.0526 compared to the initial 1.872 and 0.0493), indicating that previously concluded asymmetry might have been a false positive. Interpretation of results for other parameters remained the same.

## 4. Discussion

There is a need to identify early indicators of the risk of critical patients progressing to ICU at COVID-19. Abnormal coagulation parameters, including DD and FIB levels, changes in PLT, and PT, are observed in many patients with COVID-19 infection at admission (rev. Levi et al., 2020; Iba et al., 2020). However, as discussed by different authors, these indicators provide contradictory information regarding risk stratification and prognosis (Luo et al., 2020; Tang et al., 2020; Ding et al., 2021). There are several previous meta-analysis reviews researching abnormal coagulation and the severity of COVID-2019 infection. The majority of analyzed studies were conducted at the beginning of the pandemic and examined the Chinese population of patients (before April 2020 - Xiong et al., 2020; Lippi et al., 2020; Zhang et al., 2020; Zhu et al., 2021; June 2020 -Di Minno et al., 2020). Through our meta-analysis study, it was revealed that DD and FIB levels, APTC, and prothrombin time were significantly higher in the severe group of COVID-19 patients. In contrast, PLT was significantly lower in these patients. There was no evidence of effects across different geographic subgroups, with the exception of APTT values.

### 4.1 Platelet changes

In this meta-analysis, we systematically analyze evidence for the utility of lowered PLT levels on admission in the relationship to the severity of infection. Platelets play a crucial role in the maintenance of hemostasis, contribute to thromboinflammatory processes (van der Meijden, Heemskerk, 2019) and at different stages of viral infection, changes in platelet production or destruction may result in coagulation imbalances leading to pro-thrombotic events or in platelet disfunction and bleeding risks (Assinger, 2014), and increased activation of platelets (Hottz et al., 2020; Zaid et al., 2020; Althaus et al., 2021). Thrombocytopenia (platelet count <150×10^9^/L) seems to represent an important marker of COVID-19 severity (Qu et al., 2020; Yang et al., 2020; Lippi et al., 2020; Iba et al., 2020) and mortality outcome (Lippi et al., 2020; Yang et al., 2020). The reasons for a thrombocytopenia are multi-factorial, which may include early suppression of megakaryocytes production and damage in bone marrow and lungs, hemophagocytosis, immune destruction of platelets and other reasons. Significant thrombocytopenia in COVID-19 is less frequent than it was reported for the largest SARS-1 cohort (45%) (Lee et al., 2003). SARS-1 induced platelet depletion was related to direct infection of megakaryocytes and hematopoietic progenitors (rev. by Yang et al., 2005). However, almost half of patients presented thrombocytosis following initial thrombocytopenia during SARS epidemics in 2003 (Wong et al., 2003). Recently, a different trend in platelet changes in COVID-19 patients with immune thrombocytic purpura (ITP) was reported by Cruz-Benito et al. (2021). Secondary to SARS-CoV2 infection in ITP-patients generally develops early thrombocytosis (Cruz-Benito et al., 2020). Only a few cases of thrombocytosis (in children) were reported during SARS-CoV2 pandemics (Feld et al., 2020) as part of a multisystem inflammatory syndrome in children associated with COVID-19 (MIS-C) (Yasuhara et al., 2020). It is possible that opposite PLT changes may reflect different phases of COVID-19 infection (Thachil et al., 2020) – acute and convalescent phases, though the explanation has not been established yet. The reactive thrombocytosis that may occur during the convalescent phase requires time observation that would include a period preceding patient discharge. The increase of immature reticulated platelets reported recently in COVID-19 patients by different groups (Cohen et al., 2021; Welder et al., 2021) may play a role in the limiting of effectiveness of anti-platelet therapies (Guthikonda et al., 2007; Ibrahim et al., 2012; Armstrong et al., 2017). Change in platelets parameters (increased mean volume) (Lippi et al., 2021) and reactivity are suggested to be associated with a severe COVID-19 infection (Hottz et al., 2020; Yatim et al., 2021).

### 4.2 DD and FIB levels, and Thromboembolic complications

Concordant with other studies, we found DD and FIB levels significantly elevated in severe COVID-19 infection, though DD levels results may suffer from some publication bias as revealed by meta-regression analysis. It may be related to the variability of DD assays used in the different studies: they are not directly comparable due to the difference in the monoclonal antibodies used for tests (Olson et al., 2013; rev. Goodwin et al., 2017). As confirmed by autopsy results, the elevated DD levels can be associated with fibrin deposits within pulmonary extravascular space and alveoli (Fox et al., 2020), but they may be nonspecific to intravascular fibrin formation (Hunt et al., 2020). High DD levels, together with elevated neutrophils, were reported to be predictive factors of pulmonary embolism in COVID-19 patients (Thoreu et al., 2021). Thromboembolic complications related to COVID-19 are responsible for a substantial mortality rate (Lodigiani et al., 2020; Kunutsor, Laukannen, 2020; Piazza et al., 2020; Grillet et al., 2020). Whereas the highest burden of thromboembolism is associated with patients in the ICU (Helms et al., 2020; Llitjos et al., 2020; Klok et al., 2020), patients in non-ICU settings also bear a significant risk of these complications, especially VTE (venous thromboembolism) (Lodigiani et al., 2020; Spyropoulos et al., 2020), as well as pulmonary embolism (Thoreau et al., 2021). Klock and colleagues (2020) report that in spite of thrombosis prophylaxis, a high incidence of thrombotic complications in ICU patients is comparable with patients from other groups with DIC (disseminated intravascular coagulation). Moreover, multiple studies describe cerebral venous thrombosis (CVT) associated with COVID-19 infection (Garaci et al., 2020; Baudar et al., 2020). DD and FIB levels are elevated in COVID-19 and in thromboembolism, and therefore as single tests are unspecific and unhelpful in the differentiation of these conditions. In part, it may be explained by the involvement in abnormal coagulation at COVID-19 of different systems, including endothelial cells, complement activation, and hypofibrinolysis resulting in changes undetected by routine tests (Holter et al., 2020; Cugno et al., 2021). The dysfunction of endothelial cells with COVID-19 may contribute to excess thrombin generation and fibrinolysis shutdown, which is associated with VTE, stroke and, renal failure (Wright et al., 2020). There are different suggestions for COVID-19 risk stratification related to changes in platelets and endothelial cells, such as increased proportion of immature reticulated platelet fraction (Cohen et al., 2021; Welder et al., 2021) or CECs (circulating endothelial cells) counts in the blood representing stressed cells detached from injured vessels, reported in various inflammatory and infective diseases, and are indicative of disease severity (Blann et al., 2006). They were significantly higher in COVID-19 patients admitted to the ICU and positively correlated with the length of hospital stay (Guervilly et al., 2020). This marker is related to SARS-CoV2 directly infecting primary endothelial cells (Monteil et al., 2020). Furthermore, there is some evidence of infection in endothelial cells in severe cases of COVID-19 (Varga et al., 2020), including the presence of coronavirus particles in the cytoplasm of endothelial cells on electron microscopy (Colmenero et al., 2020). Platelets activation and endothelial cell damage in COVID-19 patients are also resulting in elevated levels of extracellular vesicles (EVs) (Capellano et al., 2021) recently associated with severity of disease (Krishnamachary et al., 2021).

### 4.3 Fibrinolysis shutdown requires evaluation outside conventional coagulation parameters

Fibrinolysis shutdown is induced by severe COVID-19 infection (Tang et al., 2020; Wright et al., 2020; Collett et al., 2021) and is partly responsible for high DD and FIB levels. Some pathophysiological changes may contribute to high procoagulant conditions secondary to COVID-19 infection. Thus, low plasminogen levels (Correa et al., 2020), similar to plasminogen-related fibrinolysis observed at other viral infections such as influenza infection (Berri et al., 2013), may lead to a hypercoagulability state. A multiplexed analysis of inflammation-related gene expression (249 genes from nCounter Inflammation panel; NanoString Technologies, USA) revealed similarities between lung specimens in influenza and Covid-19 groups (Ackermann et al., 2020). Depression of fibrinolysis was reported not only in ICU patients with severe infection and clinical signs of thromboembolism (Wright et al., 2020), but also in the general unselected ICU cohort (Correa et al., 2020; Collett et al., 2020; Nougier et al., 2020). Viscoelastic testing (ROTEM, tissue-type plasminogen activator (tPA) ROTEM, TEG) and evaluation of clot formation by Clot Waveworm analysis have demonstrated a hypercoagulable state, characterized by increased clot stiffness and severely impaired fibrinolysis (Nougier et al., 2020; Collett et al., 2020; Panigada et al., 2020; Pavoni et al., 2020; Fan et al., 2021; Hulshof et al., 2021). Elevated routine coagulation parameters taken at admission may help with initial differentiation of severe and non-severe COVID-19 patient groups, but they do not provide information about fibrinolysis shutdown.

Although our findings were consistent across different geographic groups, they were tempered by significant heterogeneity. The DD, FIB, and PLT elevation trends speak about the necessity of investigating the potential of additional specific biomarkers for stratification of COVID-19 patients at risk for fibrinolytic shutdown and associated micro-and macro-vascular events. We suggest that thorough characterization of platelets including a proportion of immature reticulated platelets and their size/mean volume parameters as well as a clot formation testing requires to evaluate a risk of thromboembolic events in COVID-19 patients groups.

## 5 Limitations of the study

Our findings should be interpreted with some limitations. Firstly, most of the included studies were either retrospective and observational by design, thus prone to recall or misclassification bias. Second, such studies often imply recruitment by convenience sampling, in which case the representativeness of a study is questioned. The random effects model was used due to the high heterogeneity of research. In the context of this meta-analysis, several different endpoints were allowed: severity guidelines, ICU admissions, oxygen therapy requirement, and disease aggravation. In an attempt to overcome this issue, we have conducted a subgroup analysis. Third, uncontrolled variables might also pose a limitation - both the disease severity and coagulation parameters can be affected by several potential confounding variables, such as comorbidities, age, etc. Some studies have reported their results in terms of quartiles or maximum and minimum values. The necessity to convert values could potentially influence the results. Although sensitivity and subgroup analyses were conducted, some residual heterogeneity may affect the interpretation of results. Finally, the underlying mechanisms of coagulation markers at COVID-19 still need to be investigated.

## 6 Conclusions

Our findings support using a combination of coagulation parameters for risk stratification of patients with COVID-19 infection at the time of admission. However, additional evaluation using clotting formation methods or combination with platelet-specific biomarkers requires. This information may help physicians triage patients with thromboembolism. An important consideration is an intermediate and long follow-up of COVID-19 patients. We hypothesize that increase in platelets turnover towards immature reticulated forms contribute to hypercoagulability state at COVID-19 patients and their resistance to anti-platelet therapy. Future research should take in account change in platelet parameters and platelet fragmentation leading to elevated levels of extracellular vesicles.

## Data Availability

All data produced in the present study are available upon reasonable request to the authors

## 7 Data availability Statement

Data are available by request from co-corresponding authors.

## 9 Author Contributions

P.L. - conceptualization, methodology, formal analysis, investigation, data curation, writing original draft, G.I., Z.S., A.K. – methodology, data curation, formal analysis, investigation; A.T.T., M.M.S., A.S.D., M.M.A., M.S.B. - methodology, formal analysis, investigation, data curation of NRCSC cohort, writing a draft; E.D.P., A.T.-methodology; analysis; funding, N.S.B. - conceptualisation, methodology, formal analysis, investigation, data curation, writing a draft. M.S.B. and N.S.B.-supervision. All authors read and edit a final draft.

## 10 Acknowledgments

We are very grateful to Sadyk Khamitov and Aliya Sailybayeva from the Research Department of National Research Center for Cardiac Surgery for their excellent organizational help. N.S.B. and A.T. were supported by FDCRGP SSH2020028, and N.S.B. by OPCRP2020018 grants from Nazarbayev University.

## 11 Abbreviations

APTT: activated partial thromboplastin time
CI: confidence interval
DD: D-dimer
DIC: disseminated intravascular coagulation
FIB: fibrinogen
GOSH: graphical display of study heterogeneity
ICU: intensive care unit
MERS-CoV: Middle East respiratory syndrome coronavirus (MERS-CoV)
NRCSC: National Research Center for Cardiac Surgery
PLT: platelets
PT: prothrombin time
ROTEM: rotation thromboelastometry
SARS-CoV: severe respiratory acute syndrome coronavirus (SARS-CoV)
SMD: standardized mean difference
tPA: tissue-type plasminogen activator
VTE: venous thromboembolism
vWF: von Willebrand factor

## Legends to Supplementary Materials

**Supplementary File 1**. Criteria for clinical severity of COVID-19 used by the National Research Center for Cardiac Surgery.

**Supplementary Table 1**. Results of the NIH Quality Assessment.

**Supplementary Table 2**. Results of the Egger’s test with Pustejovsky’s corrected standard error.

**Supplementary Figure 1**. Exploring influential cases in meta-analysis models with Baujat plots: (A) Platelets, (B) D-dimers, (C) Fibrinogen, (D) Activated partial thromboplastin time, (E) Prothrombin time.

**Supplementary Figure 2**. Influence diagnostic of meta-analysis model for the association of platelets count and COVID-19 severity.

**Supplementary Figure 3**. Influence diagnostic of meta-analysis model for the association of D-dimers concentration and COVID-19 severity.

**Supplementary Figure 4**. Influence diagnostic of meta-analysis model for the association of fibrinogen and COVID-19 severity.

**Supplementary Figure 5**. Influence diagnostic of meta-analysis model for the association of activated partial thromboplastin time and COVID-19 severity.

**Supplementary Figure 6**. Influence diagnostic of meta-analysis model for the association of prothrombin time and COVID-19 severity.

**Supplementary Figure 7**. Search for influential cases by leave-one-out method in meta-analysis models for the association of (A) platelets count, (B) D-dimers and COVID-19 severity.

**Supplementary Figure 8**. Search for influential cases by leave-one-out method in meta-analysis models for the association of (A) fibrinogen, (B) APTT and COVID-19 severity.

**Supplementary Figure 9**. Search for influential cases by leave-one-out method in meta-analysis model for the association of prothrombin time and COVID-19 severity.

**Supplementary Figure 10**. Forest plots of association of COVID-19 severity and (A) platelets count and (B) D-dimers with outliers removed.

**Supplementary Figure 11**. Forest plots of association of COVID-19 severity and (A) fibrinogen, (B) activated partial thromboplastin time, and (C) prothrombin time with outliers removed.

**Supplementary Figure 12**. Exploring the influence of meta-analysis study composition on heterogeneity and pooled effect size: (A) fibrinogen, (B) activated partial thromboplastin time, and (C) prothrombin time.

**Supplementary Figure 13**. Results of three clustering methods (A) K-means, (B) DBSCAN, and (C) Gaussian Mixture on the meta-analysis model for association of fibrinogen and COVID-19 severity.

**Supplementary Figure 14**. Results of three clustering methods (A) K-means, (B) DBSCAN, and (C) Gaussian Mixture on the meta-analysis model for association of activated partial thromboplastin time and COVID-19 severity.

**Supplementary Figure 15**. Results of three clustering methods (A) K-means, (B) DBSCAN, and (C) Gaussian Mixture on the meta-analysis model for association of prothrombin time and COVID-19 severity.

**Supplementary Figure 16**. Exploring presence of publication bias with corrected funnel plots: SMD plotted against sample size-based precision estimate: (A) platelets, (B) D-dimers, (C) fibrinogen, (D) activated partial thromboplastin time, (E) prothrombin time.

