## Supplemental data 1 for "Meta-analysis of coagulation disbalances in COVID-19: 41 studies and 17601 patients"

### *Supplementary Material 1*

**Supplementary File 1.** Criteria for clinical severity of COVID-19 used by the National Research Center for Cardiac Surgery.

- mild - fever <39°C, $SpO_2$ > 95% on room air, no dyspnea, respiratory rate < 20 breaths/min, no imaging findings of pneumonia;
- moderate - fever >38°C, $SpO_2$ 94- 95% on room air, dyspnea on exertion, respiratory rate < 20-22 breaths/min, CT extent of lung damage <50%;
- severe – any fever, $SpO_2$ 90-93%, dyspnea at less-than-ordinary activity or rest, respiratory rate 23-30 breaths/min, CT extent of lung damage >50%;
- critical - any fever, $SpO_2$ <90%, dyspnea at rest, respiratory rate >30 breaths/min, CT extent of lung damage 75-100%;

**Supplementary Table 1.** Results of the NIH Quality Assessment.

| | 1. Was the research question or objective in this paper clearly stated? | 2. Was the study population clearly specified and defined? | 3. Was the participation rate of eligible persons at least 50%? | 4. Were all the subjects selected or recruited from the same or similar populations ts? | 5. Was a sample size justification, power description, or variance and effect estimates provided? | 6. For the analyses in this paper, were the exposure(s) of interest measured prior to the outcome(s) being measured? | 7. Was the timeframe sufficient so that one could reasonably expect to see an association between exposure and outcome if it existed? | 8. For exposures that can vary in amount or level, did the study examine different levels of the exposure as related to the outcome? | 9. Were the exposure measures (independent variables) clearly defined, valid, reliable, and implemented consistently across all study participants? | 10. Was the exposure(s) assessed more than once over time? | 11. Were the outcome measures (dependent variables) clearly defined, valid, reliable, and implemented consistently across all study participants? | 12. Were the outcome assessors blinded to the exposure status of participants? | 13. Was loss to follow-up after baseline 20% or less? | 14. Were key potential confounding variables measured and adjusted statistically for their impact on the relationship between exposure(s) and outcome(s)? | Score |
|---|---|---|---|---|---|---|---|---|---|---|---|---|---|---|---|
| Liao et al. | ✓ | ✓ | ✓ | ✓ | × | NA | NA | ✓ | ✓ | NA | ✓ | NA | NA | × | 7 |
| Shang et al. | ✓ | ✓ | ✓ | ✓ | × | NA | NA | ✓ | ✓ | NA | ✓ | NA | NA | × | 7 |
| Rauch et al. | ✓ | ✓ | ✓ | ✓ | × | NA | ✓ | NA | ✓ | ✓ | ✓ | NA | NA | × | 8 |
| Gerotziafas et al. | ✓ | ✓ | ✓ | ✓ | × | ✓ | ✓ | NA | ✓ | ✓ | ✓ | NA | NA | × | 9 |
| Cen et al. | ✓ | ✓ | NR | ✓ | × | NA | ✓ | ✓ | ✓ | ✓ | ✓ | NA | NA | × | 8 |
| P. Wang et al. | ✓ | ✓ | ✓ | ✓ | × | NA | NA | ✓ | ✓ | ✓ | ✓ | NA | NA | × | 8 |
| Di Micco et al. | ✓ | ✓ | ✓ | ✓ | × | NA | NA | NA | ✓ | NA | ✓ | NA | NA | × | 6 |
| Bauer et al. | ✓ | ✓ | CD | ✓ | × | NA | ✓ | NA | ✓ | ✓ | ✓ | NA | NA | × | 7 |
| J. Zhang et al. | ✓ | ✓ | ✓ | ✓ | × | NA | NA | NA | ✓ | NA | ✓ | NA | NA | × | 6 |

| | | | | | | | | | | | | | | |
|---|---|---|---|---|---|---|---|---|---|---|---|---|---|---|
| Lopez-Castaneda et al. | ✓ | ✓ | ✓ | ✓ | × | NA | NA | NA | ✓ | NA | ✓ | NA | NA | × | 6 |
| Yue et al. | ✓ | ✓ | ✓ | ✓ | × | NA | NA | NA | ✓ | NA | ✓ | NA | NA | × | 6 |
| White et al. | ✓ | ✓ | NR | ✓ | × | NA | NA | ✓ | ✓ | NA | ✓ | NA | NA | × | 6 |
| Noh et al. | ✓ | ✓ | NA | ✓ | ✓ | NA | ✓ | ✓ | ✓ | × | ✓ | NA | NA | ✓ | 9 |
| Bergantini et al. | ✓ | ✓ | NA | ✓ | × | NA | NA | NA | ✓ | NA | ✓ | NA | NA | × | 5 |
| Jie Liu et al. | ✓ | ✓ | NR | NR | × | NA | ✓ | ✓ | ✓ | ✓ | ✓ | NA | NA | ✓ | 8 |
| Jiac Liu et al. | ✓ | ✓ | NR | NR | × | NA | ✓ | ✓ | ✓ | NR | ✓ | NA | NA | ✓ | 7 |
| J. Zhao et al. | ✓ | ✓ | ✓ | ✓ | × | NA | NA | ✓ | ✓ | NR | ✓ | NA | NA | ✓ | 8 |
| Karakoyun et al. | ✓ | ✓ | ✓ | ✓ | × | NA | ✓ | ✓ | ✓ | ✓ | ✓ | NA | NA | × | 9 |
| X. Zheng et al. | ✓ | ✓ | NA | ✓ | × | NA | ✓ | ✓ | ✓ | ✓ | ✓ | ✓ | NA | ✓ | 10 |
| Y. Zhao et al. | ✓ | ✓ | NA | ✓ | × | NA | NA | ✓ | ✓ | NA | ✓ | NA | NA | × | 6 |
| Bastug et al. | ✓ | ✓ | NA | ✓ | × | NA | ✓ | NA | ✓ | ✓ | ✓ | NA | NA | × | 7 |
| W Liu et al. | ✓ | ✓ | NA | ✓ | × | NA | ✓ | ✓ | ✓ | ✓ | ✓ | NA | NA | ✓ | 9 |
| Fu et al. | ✓ | ✓ | NA | ✓ | × | NA | NA | ✓ | ✓ | NA | ✓ | NA | NA | × | 6 |
| H. Zhang et al. | ✓ | ✓ | NR | NR | × | NA | NA | ✓ | ✓ | NA | ✓ | NA | NA | ✓ | 6 |
| Aloisio et al. | ✓ | ✓ | NA | ✓ | × | ✓ | ✓ | NA | NA | ✓ | ✓ | NA | NA | × | 7 |
| Mikami et al. | ✓ | ✓ | NA | ✓ | × | NA | NA | ✓ | ✓ | NA | ✓ | NA | NA | × | 6 |
| Young et al. | ✓ | ✓ | ✓ | NR | × | NA | NA | NA | ✓ | ✓ | ✓ | NA | NA | ✓ | 7 |
| Higuera-de-la-Tijera et al. | ✓ | ✓ | ✓ | ✓ | ✓ | NA | NA | ✓ | ✓ | NR | ✓ | NA | NA | × | 8 |

| | | | | | | | | | | | | | | | |
|---|---|---|---|---|---|---|---|---|---|---|---|---|---|---|---|
| Cugno et al. | √ | √ | CD | √ | √ | NA | NA | CD | √ | NR | √ | NA | NA | × | 6 |
| N Chen et al. | √ | √ | √ | √ | × | NA | NA | √ | √ | × | √ | NA | NA | √ | 8 |
| Wang et al. | √ | √ | × | NR | × | NA | NA | √ | √ | NR | √ | NA | NA | × | 5 |
| Long et al. | √ | √ | CD | √ | × | NA | NA | √ | √ | NR | √ | NA | NA | × | 6 |
| Yu et al. | √ | √ | √ | NR | CD | NA | NA | √ | √ | NR | √ | NA | NA | √ | 7 |
| Suleyman et al. | √ | √ | CD | √ | × | NA | NA | √ | √ | NR | √ | NA | NA | √ | 7 |
| Z. Chen et al. | √ | √ | √ | √ | × | NA | NA | √ | √ | NR | √ | NA | NA | √ | 8 |
| Ding et al. | √ | √ | √ | NR | × | NA | NA | √ | √ | NR | √ | NA | NA | × | 6 |
| Q. Chen et al. | √ | √ | √ | × | × | NA | NA | √ | √ | NR | √ | NA | NA | × | 6 |
| G. Zhang et al. | √ | √ | √ | NR | × | NA | NA | √ | √ | NR | √ | NA | NA | × | 6 |
| Jiao Liu et al. | √ | √ | √ | √ | × | √ | NA | √ | CD | √ | √ | NA | NA | √ | 9 |
| Zhou et al. | √ | √ | √ | √ | × | √ | √ | √ | √ | NR | √ | NA | NA | √ | 10 |

**Supplementary Table 2**. Results of the Egger's test with Pustejovsky's corrected standard error

| Parameter | Intercept | Confidence Interval | t | p-value |
|---|---|---|---|---|
| PLT | -1.306 | [-2.47, -0.14] | -2.195 | 0.0355 |
| DD | 3.228 | [1.24, 5.21] | 3.184 | 0.0030 |
| FIB | 2.142 | [0.1, 4.18] | 2.061 | 0.0526 |
| APTT | 0.717 | [-1.46, 2.89] | 0.647 | 0.5255 |
| PT | -0.506 | [-2.59, 1.58] | -0.476 | 0.6389 |
